# ProgNet: Covid-19 prognosis using recurrent and convolutional neural networks

**DOI:** 10.1101/2020.05.06.20092874

**Authors:** Mohamed Fakhfakh, Bassem Bouaziz, Faiez Gargouri, Lotfi Chaari

## Abstract

—Humanity is facing nowadays a dramatic pandemic episode with the Coronavirus propagation over all continents. The Covid-19 disease is still not well characterized, and many research teams all over the world are working on either therapeutic or vaccination issues. Massive testing is one of the main recommendations. In addition to laboratory tests, imagery-based tools are being widely investigated. Artificial intelligence is therefore contributing to the efforts made to face this pandemic phase.

Regarding patients in hospitals, it is important to monitor the evolution of lung pathologies due to the virus. A prognosis is therefore of great interest for doctors to adapt their care strategy. In this paper, we propose a method for Covid-19 prognosis based on deep learning architectures. The proposed method is based on the combination of a convolutional and recurrent neural networks to classify multi-temporal chest X-ray images and predict the evolution of the observed lung pathology. When applied to radiological time-series, promising results are obtained with an accuracy rates higher than 92%.

## I. Introduction

Coronaviruses are a large family of viruses that can cause a variety of diseases in humans, ranging from colds to Middle East Respiratory Syndrome (MERS) and Severe Acute Respiratory Syndrome (SARS). In recent months, the World Health Organization (WHO) has declared that a new coronavirus called COVID-19 has spread aggressively to several countries around the world [1]. COVID-19 can cause respiratory tract disease, fever and cough, and severe pneumonia in some extreme cases [2]. Pneumonia is an infection that causes inflammation mainly in the air sacs of the lungs responsible for oxygen exchanges [3].

The COVID-19 pandemic can be considered serious due to its high contagion and severity [4]. The impact on healthcare systems is also high due to the number of people requiring intensive care units (ICU) admission and mechanical ventilators for long periods [5]. Making evidence of COVID-19 infection is generally associated with specific lung pathologies. The most reliable way is to perform PCR (Polymerase Chain Reaction) [6] tests to asses the presence of Covid-19 RNA (Ribonucleic Acid) in the hosting individual. Because of low testing capacity especially at the very beginning of the pandemic spread, most countries performed tests generally to persons suffering from unclear and advanced symptoms, or individuals who evidently have been in contact with infected ones.

In this sense, chest X-ray imaging is a technique that plays an important role in the diagnosis of COVID-19 disease. Radiological tests may be performed by analyzing chest X-ray images and identifying lung tissues potentially hosting infections. However, X-ray data analysis requires an expert in radiology, and may be time-consuming. Therefore, developing an automated analysis system may solve this problem and help saving precious time for radiologists. In this context, solutions based on artificial intelligence (AI) can provide an accurate and inexpensive diagnosis for COVID-19 and other types of pneumonia [7,8]. Furthermore, it is important to assess the pathology evolution for infected people, especially those suffering from severe lung pathology who may need intensive care. This is mainly essential since no specific treatment is known up to date, which means that a treatment line adaptation may be recommended if no improvement is observed. Pathology prognosis is also useful for prioritization of patients to take in charge in case of limited numbers of ICUs.

In this paper, we propose a method for pathology prognosis based on analysing time series of chest X-ray images. The proposed method is based on both recurrent and convolutional neural networks, and allows to classify patients into two severity classes: positive or negative evolution. The main originality lies in the use of such a combination for COVID-19 prognosis.

The rest of this paper is organized as follows. After an introduction covering the context of this research and the problem to be handled, Section II is devoted to a state of the art linked to our research. In Section III, an overview of the proposed approach is presented and all the steps are detailed for the multi-temporal classification of X-ray images. An experimental validation is illustrated in Section IV. Finally, conclusions and future work are drawn in Section V.

## II. Related work

Time series data refer to a consistent flow of data sets over a period of time. The analysis of these series has become a recent area of interest in artificial intelligence. Accurate forecasting is becoming more and more vital in all areas in order to make more informed and precise decisions. Time series analysis is mainly used for: *i)* descriptive analysis, i e, identifying trends in the correlated data, *ii)* forecasting, to predict short-term events and *iii)* intervention analysis to see how the event can be evaluated during the time series.

On the other hand, analysing time series generally presents the drawback related to the lack of annotated data sets. Annotations for time *T_n_* generally have to account for times *T_i_* with *i < n*. Having comparable data (same quality, size,…) is therefore an additional problem that leads to a difficulty in designing specific algorithms. In this sense, several applications have addressed this problem especially in clustering [9], change classification [10], change detection [11,12], and forecasting [13].

In medical imaging, time series are often used for functional [14–16] and spectroscopic [17] imaging, as well as motion analysis [18]. In this sense, X-ray images, and specifically CT (Computerized Tomography) is widely used due to its high spatial and temporal resolutions. As regards lung diagnosis, X-ray images are usually used to track pneumonia or other specific diseases.

To analyse 3*D + t* (3*D*+time) data, one usually need to design sophisticated algorithms that are automatic, user-friendly, fast and accurate. Today, Deep Learning (DL) has demonstrated its efficiency as stated hereabove in many fields of image analysis. Specifically, CNNs [19] are among the mostly used networks. The specificity of CNN lies in its capacity to automatically learn implicit features from images, in contrast to classical machine learning methods where the designer has to specify the features to be extracted and how to calculate them. Since 2012, several models of deep convolutional neural networks (DCNN) [20] have been proposed such as AlexNet [21], VGG [22], GoogleNet [23], Residual Net [24], DenseNet [25] and CapsuleNet [26].

As regards data related to Covid-19, very recent works have already started to design diagnosis aid tools based on DL in order to detect Covid-19 specificities in chest X-ray images. All the approaches used deep learning for image classification as Covid or non-Covid. In [27], a modified version of the ResNet-50 network has been proposed, on which a Feature Pyramid Network (FPN) is used to identify and automatically extract lesions from CT images. Using this module, the model can detect and classify CT images into three classes: healthy, COVID-19, and bacterial pneumonia. Likewise, chest radiographic images (CXR) have been used in [28]. The authors use a CNN based on various ImageNet^1^ pre-trained models to extract the high-level features. Those features were fed into a Support Vector Machine (SVM) as a machine learning classifier, in order to classify the radiographic images affected by the corona effect coming.

In [29], the authors use a COVID-Net architecture with transfer learning to classify CXR images into four classes: normal bacterial infection, infection non-COVID, and COVID-19 virus. Another study [30] adopts a DeTraC deep CNN architecture [31] where the general idea is to add a class decomposition layer to the preformed models. The class decomposition layer aims to partition each class of the image dataset into several sub-classes. New labels are then assigned to the new set, where each subset is considered as an independent class, then these subsets are assembled to produce the final predictions.

Despite the efficiency and the performances obtained by the various techniques described above for the detection of COVID-19, these approaches do not account for temporal correlations and only provide a decision about a single image. Prognosis remains therefore an open issue. Indeed, once a patient is declared infected, especially for those taken in charge in hospitals, it is important to monitor the evolution of the lung pathology. This monitoring can be done by checking blood oxygen level or other metrics. However, an accurate prognosis has mainly to be done by controlling lung images. In this paper, we propose a method to analyse time series of lung images related to infected patients in order to derive a prognosis whether the pathological state is getting better or not.

Recurrent neural networks (RNN) are a family of deep learning methods designed to manage temporal correlations between images in time series. These networks have recurrent connections in the sense that they keep information in memory: they can take into account at time *T_n_* a number of past states *T_i_* where *i < n*. These networks have been used in remote sensing to assess change detection in multi-spectral and hyper-spectral images [32]. RNNs are able to memorize information for a limited time and start to ”forget” after a certain number of iterations, which makes training for many applications complicated. The algorithms used for updating the weight in RNNs are mainly based on the gradient with some well known practical problems such as gradient explosion.

To overcome this limitation, Long-Term Short-Term Memory (LSTM) have been proposed in [33–35] as a particular type of RNN. These models explicitly capture recursive temporal correlations and they have already proven their effectiveness in various fields, such as speech recognition [36], natural language processing [37] and image completion [38]. LSTMs have recently been used in medical imaging such in [39] where the authors propose a method with multi-modality and adjacency constraints for the segmentation of the cerebral image.

Finally, it is worth noting that LSTMs and CNNs have already been combined in a number of works like [40–42]

## III. Methodology

### A. General overview

The proposed methodology consists of applying a deep learning architecture for the multi-temporal classification of X-ray images in order to evaluate the COVID-19 evolution, and hence draw a vital prognosis for infected patients.

The proposed methodology is based on the combination of RNN and CNN architectures in order to assess the temporal evolution of images, and hence classify the time series into two main classes: positive and negative evolution. Such a classification can help doctors guess a vital prognosis for patients in critical situations.

Our architecture is capable of automatically learn temporal correlations of images provided as input. Fig. 1 gives a general overview of the proposed ProgNet architecture.

**Fig. 1.**
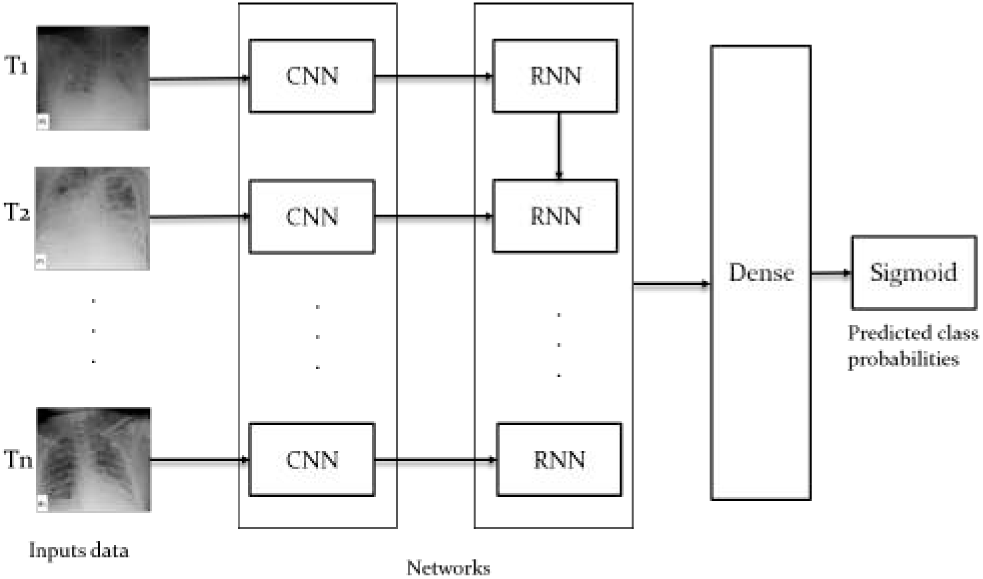
Overview of the proposed ProgNet architecture.

For an X-ray sequence *T* = (*T*_1_, *T*_2_*,…, T_n_*), each image *T_i_* goes through a CNN in order to extract a characteristic vector. The obtained vector involves a set of information subsequently generated by applying several convolutions and pooling layers. As a specific configuration, ResNet50 is one of the potential candidates to be used as a CNN which has shown its efficiency in different image classification applications.

An RNN is then applied to learn temporal correlations between the different characteristic vectors related to each image of a sequence *T*. LSTM is used due to its good performance demonstrated in the literature [33–35]. Four fully connected layer with a *sigmoid* decision function are then applied to perform binary classification.

A detailed description of the adopted networks is provided in Section III-B.

### B. Used networks

#### 1) ResNet

Residual networks (ResNet) is a classic CNN used as a backbone for many computer vision tasks. This model won the ImageNet challenge in 2015. In deep networks, low, medium, and high functionality and classifiers are extracted into a set of layers. ResNet mainly solves two key problems generally faced in the training deep neural networks: vanishing and exploding gradients. The core idea of ResNet is to introduce the ’’identity shortcut connection” which skips one or more layers. Fig. 2[left] illustrates plain layers where each convolution is connected to each other, which is the idea of all architectures before the appearance of ResNet, while Fig. 2[right] shows the residual network (Skip connection). Skipping layers allows avoiding gradient to vanish during back-propagation. Using such connections, one can train extremely deep networks that can take advantage of the power of depth, and hence allow capturing complex patterns in data.

**Fig. 2.**
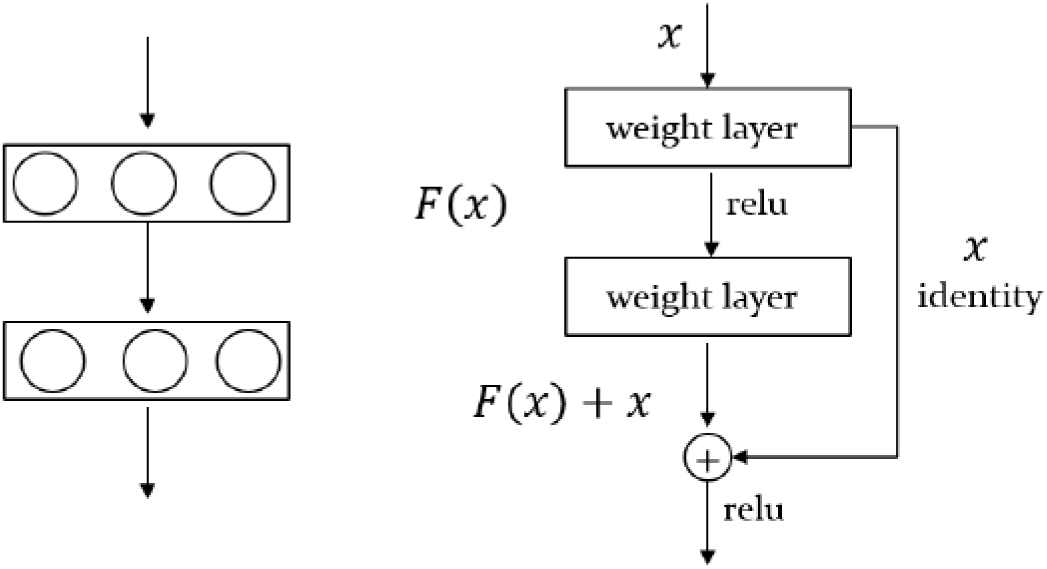
Plain layers (left) and Residual blocks with skip connection (right).

As illustrated in Fig. 3, the ResNet50 [24] architecture used in this study is made up of 5 blocks, where each of them contains a set of convolution and max-pooling layers, followed by a skip connection.

**Fig. 3.**
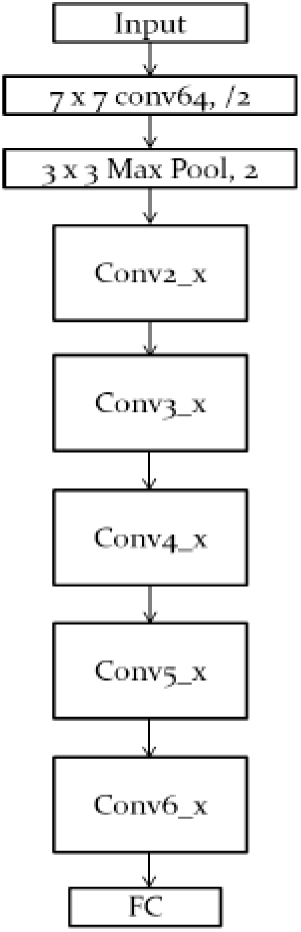
Summarised overview of the ResNet50 architecture.

#### 2) LSTM

In some use cases, it is important to know what decisions have been made in the past in order to make an optimal decision at time t. LSTMs are adopted here since mixing long and short term dependencies in chest X-ray images is important and complementary with spatial dependencies analyzed by our CNN. LSTMs have sophisticated dynamics that allow it to easily “memorize” information for an extended number of time-steps. The ’long term” memory is stored in a vector of memory cells {*C*_1_, *C*_2_,…*C_n_* }.

Although some differences exist in LSTM architectures, all of them have explicit memory cells for storing information for long periods of time. The LSTM can decide to overwrite the memory cell, retrieve it, or keep it for the next time step. A typical LSTM cell is illustrated in Fig. 4.

**Fig. 4.**
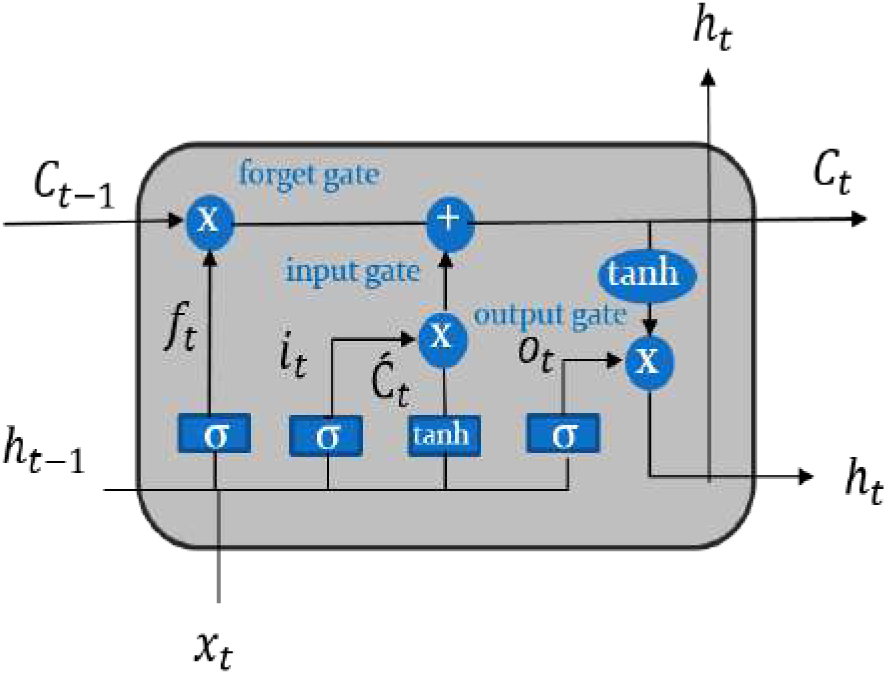
Detailed LSTM cell architecture.

In Fig. 4, the cell input is denoted by *x_t_* (input data), while *h_t_*_−1_ stands for the state hidden at the previous step, and *C_t_*_−1_ for the previous memory cell. The cell outputs are the next hidden state *h_t_* and memory cell *C_t_*.

Two main points distinguish an LSTM cell from a standard recurrent layer. First, the cell state is split into two parts, the long-term *C_t_* and short-term *h_t_* states. Second, three control gates along the state path (forget, input, and output gates) are added to regulate the cell states.

The first step of an LSTM cell is to decide what information will be thrown out of the cell state. The decision is made by a *sigmoid* layer called ’forget gate layer (*f_t_*)”. It evaluates *h_t_*_−1_ and *x_t_* to generate a number between 0 and 1 for each cell state *C_t_*_−1_ coefficient. The value 1 represents ’’completely keep this” while 0 represents ’completely get rid of this”. Eq.(1) shows how to control the information removal from the previous long-term state *C_t_*_−1_:

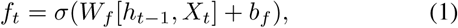

where *σ* is the *sigmoid* function, *W_f_* and *b_f_* corresponds to the weight matrix and bias.

The next step is to decide what new information has to be stored in the cell state. First, a *sigmoid* layer called the ’input gate layer *i_t_”* (Eq. (2)) decides which values to update. Next, a hyperbolic tangent *(tanh)* layer creates a vector of new candidate values 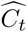 (Eq. (3)), that will be added to the state. These elements are then combined to create a state update:

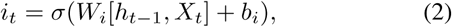

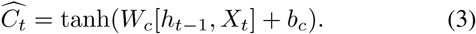

As in Eq. (1), *W_i_* and *W_c_* stand for weight matrices, while *b_i_* and *b_c_* are the bias terms.

In an LSTM cell, the old cell state *C_t_*_−1_ has then to be updated into the new cell state *C_t_* following Eq.(4):

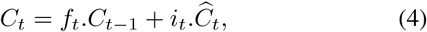

where ”.” is the point-wise matrix multiplication. The last step is to decide what information to produce. The output gate *o_t_* calculation is based on the cell state following Eq. (5), while the hidden state is updated according to Eq. (6):

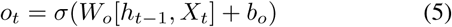

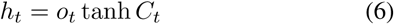

In (5), *W_o_* and *b_o_* correspond to the weight matrix and bias. The output of the block is recurrently connected back to the block input and all of the gates.

## IV. Experimental validation

In this Section, we illustrate the performance of the proposed architecture for COVID-19 prognosis. Experiments are conducted on an open database of COVID-19 cases with chest radiographs^2^. To the best of our knowledge, this is the only available dataset containing temporal acquisitions for a number of patients, with ground truth annotation as a therapeutic issue for each patient: death or survival. This database contains data for 42 patients, with up to 5 images for the same subject. For the validation of the proposed architecture, we used 34 sequences for training, each includes 3 X-ray images of the same patient, and 8 sequences for the test.

To implement our progNet architecture, we put four dense layers respectively, FC-1024, FC-500, FC-128, and FC-64. As regards coding, we used python programming language with Keras and Tensorflow libraries on a Intel(R) Core(TM) i7-3630QM CPU 2.40GHZ architecture with 8 Go memory.

In order to asses the performance of the proposed progNet architecture on the available data, we performed comparisons with three other possible configurations. Each configuration relies on a different CNN (see Fig. 1): AlexNnet, VGG16, VG19. These networks have already demonstrated an outstanding performance in the literature.

### A. Loss and accuracy behavior

We first assess and compare the training and validation errors during the training procedure.

We used an ADAM optimization technique with a learning rate of 10^−4^ and binary cross-entropy loss. The minimum batch size was 32 and 60 epochs were considered. The weight decay was set to 10^−4^ to prevent over-fitting while training the model. The momentum value was set to 0.9. As regards the depth of the used LSTM, we defined 150 hidden units and added a dropout layer of 0.5.

Fig.5 displays the obtained train accuracy and loss curves for the proposed ProgNet (Fig.5[(d)]) as well as the other used configurations: AlexNnet (Fig.5[(a)]), VGG16 (Fig.5[(b)]), VG19 (Fig.5[(c)]). The displayed curves indicate similar convergence performance for the four architectures.

**Fig. 5.**
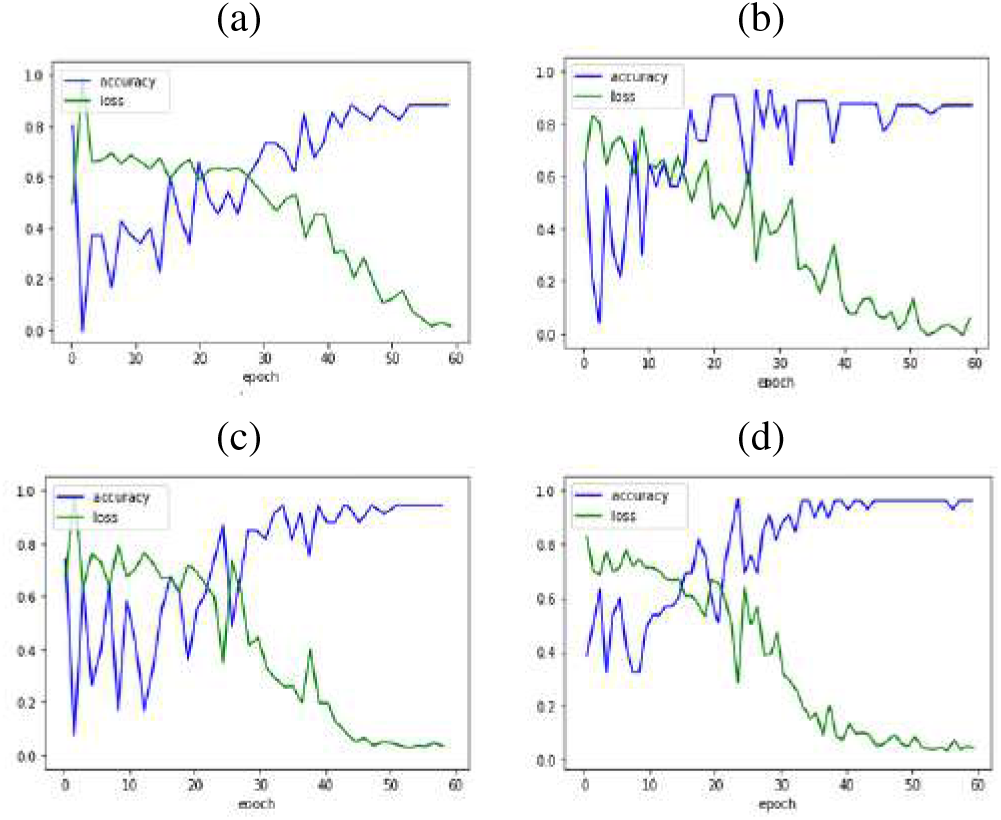
Accuracy/Loss of the learning train obtained with (a) AlexNet, (b) VGG16, (c) VGG19 and (d) ProgNet.

As regards the test set, Fig. 6 displays accuracy and loss curves for all architectures. When visually inspecting Fig. 6[(d)], we see that the loss and accuracy curves get better faster for the ProgNet architecture with a more stable behavior. Training with the ProgNet architecture is therefore faster and more efficient that the other configurations, partly due to the CNN depth.

**Fig. 6.**
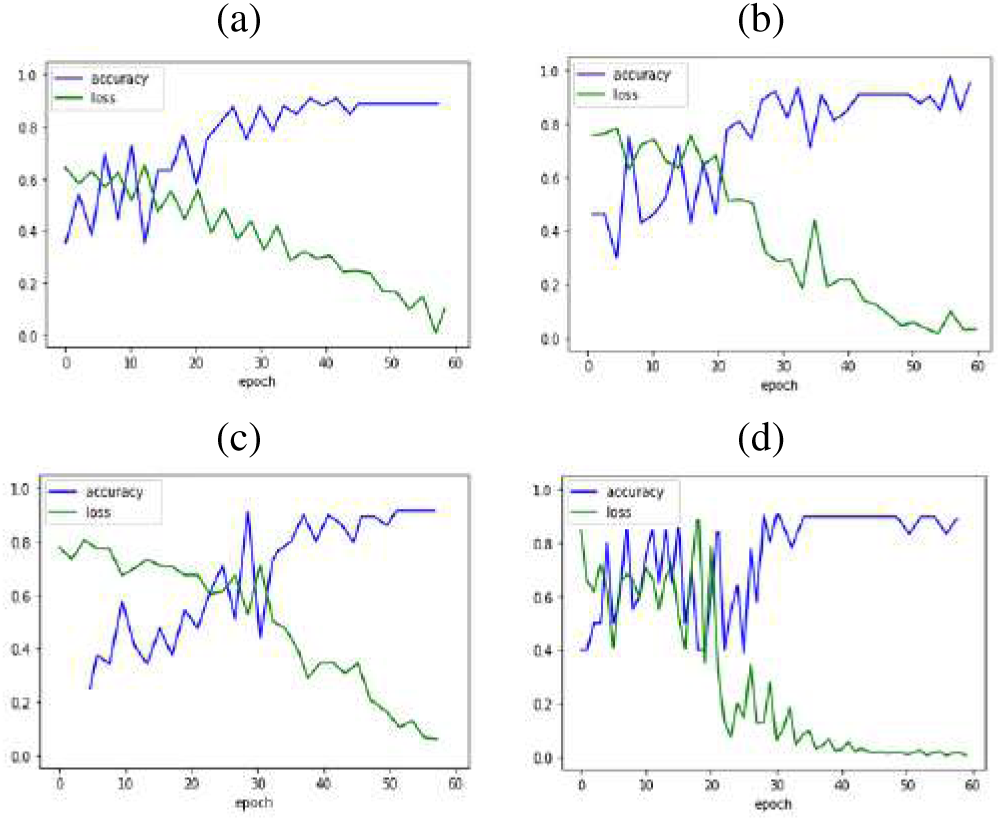
Accuracy/Loss of the learning test curves obtained with (a) AlexNet, (b) VGG16, (c) VGG19 and (d) ProgNet.

It is worth noting that the observed peaks in the accuracy and loss curves may be caused by a lack of representatives of the training and test data, and hence a difficulty in learing some fine features. This deserves more investigation to assess the ability of the model to generalize [43,44]. This is mainly due to the limited number of available time-series of X-ray chest images related to Covid-19.

### B. Quantitative evaluation

To further assess the performance of the proposed method, Table I reports obtained values for the rates of True Positives (TP), True Negatives (TN), False Positives (FP) and False Negatives (FN). As reported in Table I, all the networks perform well with TP rates over 87%. However, the proposed ProgNet architecture slightly outperforms the others with the highest TP and TN rates, while the FN and FP rates are also the lowest. By enjoying the lowest FN, the proposed architecture appears very interesting since for such a prognosis, it is crucial to not miss negative evolution cases.

**TABLE I.** TP, FN, FP and TN values for the proposed ProgNet method and AlexNet, VGG16, VGG19 architectures.

|  | TP | FN | FP | TN |
| --- | --- | --- | --- | --- |
| **AlexNet** | 88.37% | 11.96% | 11.63% | 88.04% |
| **VGG16** | 90.14% | 8.79% | 9.86% | 91.21% |
| **VGG19** | 91.90% | 7.89% | 8.1% | 92.11% |
| **ProNet** | **93.81%** | **5.99%** | **6.19%** | **94.01%** |

To further assess the quantitative performance, accuracy, precision and recall are also provided in Table II, in addition to the *F*_1_ score [45]. The *F*_1_ score takes into consideration both precision and recall in order to validate the accuracy. It is the harmonic mean of both measures and can be calculated as

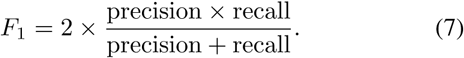

**TABLE II.** Mean accuracy, precision, recall and F1Score for the proposed ProgNet architecture as well as AlexNet, VGG16, VGG19.

|  | AlexNet | VGG16 | VGG19 | ProgNet |
| --- | --- | --- | --- | --- |
| **Accuracy** | 0.888<br>$\pm 0.08$ | 0.905<br>$\pm 0.03$ | 0.923<br>$\pm 0.04$ | **0.924**<br>$\pm 0.03$ |
| **Precision** | 0.883<br>$\pm 0.05$ | 0.90<br>$\pm 0.02$ | 0.91<br>$\pm 0.03$ | **0.930**<br>$\pm 0.02$ |
| **Recall** | 0.880<br>$\pm 0.09$ | 0.911<br>$\pm 0.05$ | 0.920<br>$\pm 0.06$ | **0.939**<br>$\pm 0.04$ |
| **$F_1$ score** | 0.881<br>$\pm 0.06$ | 0.905<br>$\pm 0.03$ | 0.914<br>$\pm 0.04$ | **0.934**<br>$\pm 0.02$ |

In Table II, the mean values over 10 runs are provided. For each run, a sub-set of training data is randomly chosen. Standard deviations over the 10 runs are also provided in the table.

Through the reported values, one can easily notice that the proposed ProgNet method outperforms the other competing architectures. Specifically, higher precision and recall values indicate that ProgNet is more efficient in retrieving ambiguous infection cases. Moreover, the reported low standard variation values show better stability for the proposed model, which indicates better generalization properties. A visual inspection of ambiguous time-series is provided in Section IV-C.

### C. Qualitative Analysis

In this section, we illustrate some representative results obtained on a group of patients. These time series are made up of 3 images acquired at times *T*_1_, *T*_2_ and *T*_3_. Fig. 7 displays four time series of covid-19 patients, two of them (top) have survived, while the two others (buttom) are dead. For each patient, the obtained ’’survival score” (SC) is provided on the first column. This score is nothing but the probability predicted by the *sigmoid* function.

**Fig. 7.**
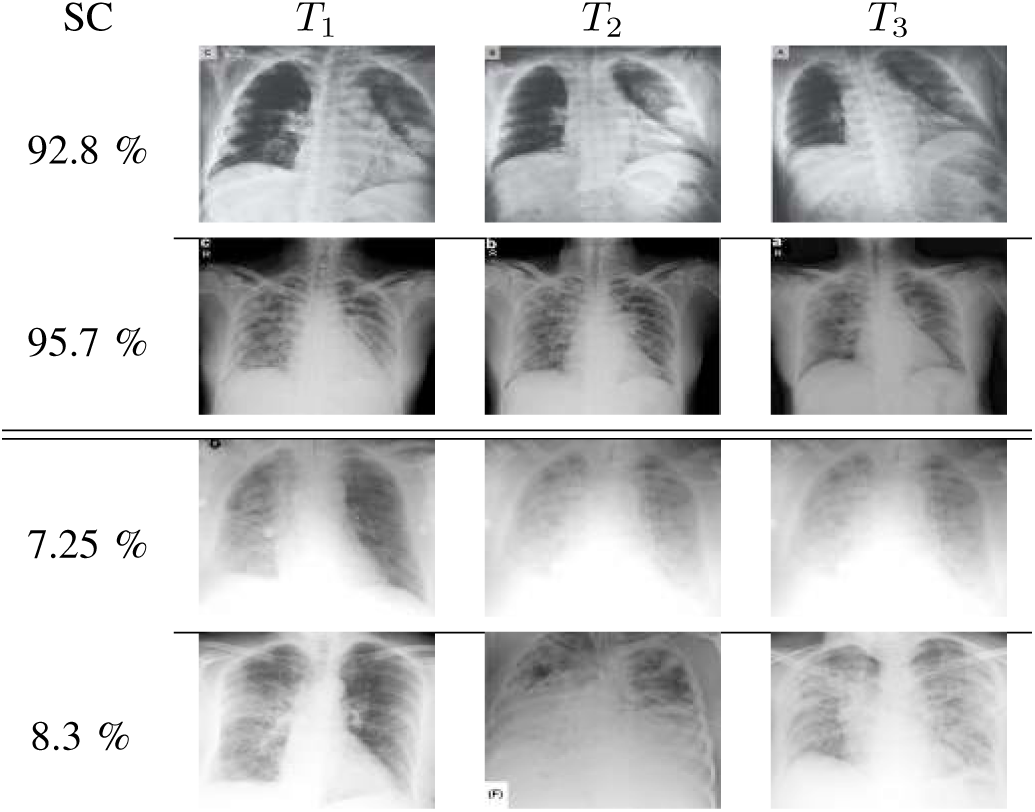
Visualization of the COVID-19 classification using the propose ProgNet architecture.

From a visual point of view, it is clear that time-series with death issue show more white areas over all images. Generally speaking, the spread of these areas increases over time. Reported SC values indicate how sure the ProgNet model is about the classification results for these time-series.

As reported in the previous section, good precision and recall values indicate the ability of our ProgNet architecture to properly classify ambiguous cases. In this sens, Fig. 8 shows two examples of time-series corresponding to two patient with a survival and death issues. These patient are well classified by the proposed ProgNet architecture, while the competing methods mis-classify them. when visually inspecting the images for the death issue, ambiguity comes from extended white areas in all the images with no visible improvement over time. The same remark holds for the survival case where white areas persist over time.

**Fig. 8.**
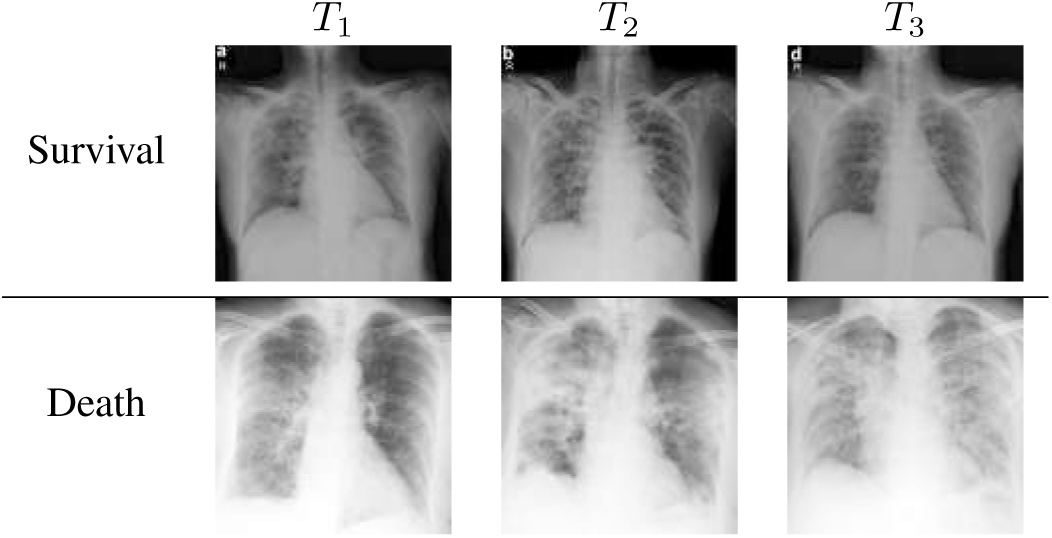
Examples of time-series well classified by ProgNet for which the competing methods fail.

To investigate why the proposed ProgNet architecture fails to classify some cases, Fig. 9 shows images of a time-series linked to a patient mis-classified by ProgNet. Indeed, we notice in some cases an unexpected change in the situation of a patient during the last image of the sequence. This can lead to some problems in learning all features and managing the time correlation between them. Analysing longer sequences can certainly improve the model performance.

**Fig. 9.**
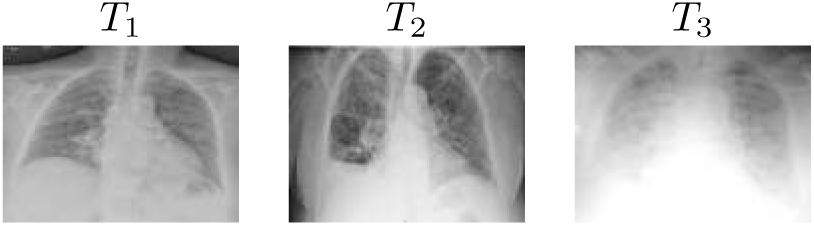
Images of an ambiguous time-series mis-classified by ProgNet.

## V. Conclusion

In this paper, we proposed an architecture for Covid-19 prognosis, based on a combination of a CNN and RNN networks. The proposed architecture analyses both spatial and temporal dependencies in input time-series of chest X-ray images. Our method is segmentation-free, which makes it competitive with respect to other assessment methods relying on time-consuming lung segmentation algorithms. when applied on radiographic data, the proposed ProgNet architecture showed promising results with good classification performances, especially for ambiguous cases. Specifically, the reported low false positive rates are interesting for an accurate and personalised care workflow.

Future work will focus on applying the proposed architecture to CT data with longer time-series. However, data availability and homogeneity is a key issue. In this sense, the proposed architecture has to be adapted in order to handle data heterogeneity in time (time-series with different sizes).

## Data Availability

data available here https://github.com/ieee8023/covid-chestxray-dataset/

## Footnotes

1 http://www.image-net.org/

2 https://github.com/ieee8023/covid-chestxray-dataset/

